# Racial disparities in COVID-19 mortality across Michigan, United States

**DOI:** 10.1101/2020.11.30.20241133

**Authors:** Alyssa S. Parpia, Abhishek Pandey, Isabel Martinez, Abdulrahman M. El-Sayed, Chad R. Wells, Lindsey Myers, Jeffrey Duncan, Jim Collins, Meagan C. Fitzpatrick, Alison P. Galvani

**Affiliations:** Center for Infectious Disease Modeling and Analysis (CIDMA), Yale School of Public Health, New Haven, CT, USA; Department of Internal Medicine, University of Michigan Medical School, Ann Arbor, MI, USA; Department of Public Health, Wayne State University, Detroit, MI, USA; Division for Vital Records and Health Statistics, Michigan Department of Health and Human Services, Detroit, MI, USA; Division of Communicable Diseases, Michigan Department of Health and Human Services, Detroit, MI, USA; University of Maryland School of Medicine, Baltimore, MD, USA

## Abstract

Black populations in the US are disproportionately affected by the COVID-19 pandemic, but the increased mortality burden after accounting for health and demographic characteristics is not well understood. We evaluated COVID-19 mortality in Michigan using individual-level death certificate and surveillance data from the Michigan Department of Health and Human Services from March 16 to October 26, 2020. Among the 6,065 COVID-19-related deaths, Black individuals experienced 3.6 times the mortality rate as White individuals. Black individuals under 65 years without comorbidities had a mortality rate 12.6 times that of their White counterparts. After accounting for age, sex, and comorbidities, we found that Black individuals in all strata are at higher risk of COVID-19 mortality than their White peers. We demonstrate that inequities in mortality are driven by ongoing systemic racism, as opposed to comorbidity burden or older age, and further highlight how underlying disparities across the race are compounded in crises.

## Introduction

Throughout history, epidemics have inequitably affected vulnerable populations in our societies,^1,2^ and the COVID-19 pandemic is no exception. In particular, Black populations are disproportionately experiencing severe COVID-19 morbidity and mortality in the United States (US).^3,4^ According to data from the Centers for Disease Control, 53% and 23% of all COVID-19 deaths in the US are among White and Black individuals, respectively, while these races represent 42.1% and 17.0% of the population.^5,6^ In Michigan, the disparities are even starker: while Black Americans represent 14.1% of the total population,^7^ 35.0% of state-wide COVID-19 deaths have occurred in this group as of November 5, 2020.^8^ Underlying this disparate burden are systemic inequities in socio-economic conditions and health by race, which impact infection exposure and survival.

Longstanding marginalization of Black communities has led to higher rates of housing instability, financial insecurity, and essential service inaccessibility compared to White communities.^9–11^ For instance, Black families in Michigan report 2.5 times higher rates of poverty^12^ and 43% lower household incomes^13^ than White families. Flint and Detroit, the residents of which are predominantly Black, are the the second and fourth poorest cities in the US, respectively.^14^ Socioeconomic disparities can influence community-level COVID-19 transmission, as smaller living space, greater household sizes, and reliance on public transportation each impact the potential risk for COVID-19 infection. As a result of disparities in access to healthcare^15^ and other aspects of structural racism, Black populations also experience higher rates of chronic health conditions such as diabetes,^16^ obesity,^17^ asthma,^18^ and cardiovascular disease^19^ in comparison to White populations, which affect the severity of disease following infection.^20^

This multitude of factors creates a challenge for individuals in general, and especially those with underlying medical conditions, to protect themselves from infection. The “stay at home” public health guidelines implemented during the COVID-19 pandemic are in tension with the need for stable employment, wages, and housing. Pandemic related job losses have been experienced predominantly by those in lower income brackets,^11^ those in Black communities,^11^ and those unable to perform work tasks at home.^21^ When a certain segment of the population is fundamentally unable to follow infection prevention guidelines, disparities in morbidity and mortality inevitably arise.

We analyzed individual-level data on people who died from COVID-19 in Michigan, stratified by demographic characteristics, underlying medical conditions, and geographical location. We assessed disparities in mortality risk by race within each ZIP code between March 16 to October 26, thereby evaluating the interdependent crises of the COVID-19 epidemic and systemic racism in Michigan. Further, we used statistical analyses to assess disparities across races among individuals dying from COVID-19 in Michigan. We found that Black individuals have 3.6 times the risk of dying from COVID-19 as White individuals in Michigan overall. Among those with no comorbidities under the age of 65, Black individuals have 12.6 times the mortality rate of White individuals.

## Methods

### Data Sources

#### Individual-level COVID-19 Surveillance and Death Certificate Data

Linked death certificate and COVID-19 surveillance data were obtained from the Michigan Department of Health and Human Services. The Division of Vital Records and Health Statistics provided death certificate data for individuals with an International Statistical Classification of Diseases and Related Health Problems, Tenth Revision (ICD-10) code for COVID-19 (U07.1 or U07.2) as an underlying or related cause of death and the Communicable Diseases Division provided data from the Michigan Disease Surveillance System on COVID-19 deaths occurring between March 16 and October 26, 2020. All COVID-19 related deaths occurred in the state of Michigan. Individual level data were collected on age, sex, race, ZIP Code Tabulation Area (hereafer referred to as ZIP code) of residence, underlying and related causes of death, other medical conditions of interest, pre-existing conditions, immunosuppressive medications, retirement or unemployment status, and residence or employment at a high risk or congregate living facility.

Comorbidities were identified by combining information on underlying causes of death, related causes of death, and other medical conditions of interest from death certificates with information on pre-existing conditions and medications from the Surveillance System case report forms. Comorbidities were categorized as follows: asthma or reactive airway disease, cardiovascular disease, cancer, chronic lung disease, diabetes mellitus, neurologic disease, chronic liver disease, chronic renal disease, other immunosuppressive conditions, and other chronic diseases. Other immunosuppressive conditions included rheumatoid arthritis and bullous pemphigoid, among others, as well as those unspecified but noted as immunosuppressive. Other chronic diseases included anemia, depression, and chronic venous thromboembolism as well as those unspecified (See Supplement Section 3 for complete lists). Cardiovascular disease corresponded to ICD-10 codes I00-I78, and includes heart attacks and strokes. Comorbidity of cancer represented any history of cancer.

ZIP code of residence was obtained primarily from death certificate data and from surveillance system case reports in instances of missing ZIP code on the death certificate. Only individuals who resided in Michigan were included in the mapping. Individuals who died from COVID-19 were categorized by their status of being residents or employees of high risk or congregate living facilities, which include: long-term care homes, skilled nursing facilities, assisted living facilities, homeless shelters, federal prisons, Michigan Department of Corrections prisons, county jail, juvenile justice facilities, foster care, and others, including senior, retirement, and group homes.

#### Population-level Demographic Data

Data on Michigan population demographics by age group, sex, and race were obtained from the Michigan Department of Health and Human Services.^22^ The prevalence of comorbidities in the Michigan population by age group, sex, and race were estimated using a combination of 2017 Michigan Medicare data^23–25^ and National Health Interview Survey data on national comorbidity distribution by age group and sex^26^ (See Supplement Section 2 for calculations).

We assumed that race did not affect the sex distribution of comordities. Populations by ZIP code for all races were obtained from the United States Census Bureau Decennial Census.^27^

### Analysis

Using a Chi-Square test, we determined whether a statistically significant difference (α = 0.05) existed between the proportions of deaths which occurred among Black and White individuals in Michigan, and the proportions of the overall population who are Black and White in Michigan.

We also performed univariate and bivariate analyses on COVID-19 mortality by demographic characteristics and presence of single or multiple comorbidities. Specifically, Chi-square and Kruskal-Wallis tests were used to identify differences in the following variables by race: age, sex, number of comorbidities, presence of specific COVID-19 related comorbidities and comorbidity combinations, employment status, and high-risk or congregate living facility exposure as resident or staff. In addition, COVID-19 mortality rates and mortality rate ratios were calculated for the overall sample, as well as by race, age, sex, and number of comorbidities both separately and in combination. Chi-square tests were performed to identify differences in these mortality rates by race.

We combined the individual-level data on COVID-19 deaths with ZIP code level data on total population and population by race to evaluate geographic heterogeneity in mortality. For each ZIP code, we calculated the COVID-19 mortality rate overall and by race, as well as the mortality rate ratio of Black to White deaths. A one-tailed Wilcoxon signed-rank test was used to compare whether the mortality rates among Black individuals were greater than those of White individuals across ZIP codes throughout Michigan.

We conducted descriptive analyses of the dates of COVID-19 symptom onset, hospitalization, and death due to COVID-19, stratified by race, to characterize epidemic progression. We then conducted an analysis of variance (ANOVA) in order to identify stratum-specific differences in the time from COVID-19 symptom onset to hospitalization, accounting for age group, sex, race, and number of comorbidities. All data were analyzed using Python 3.7.4 and R-3.5.1.

## Results

### Demographics of COVID-19 Decedents

Between March 16 and October 26, 2020, a total of 6,065 COVID-19 related deaths were recorded in the surveillance and death certificate databases together, of which 96.3% occurred among Black or White individuals. Black individuals represent 15.7% of the combined Black and White population of Michigan yet accounted for 40.1% of the COVID-19 deaths. The proportion of deaths reported among Black individuals in Michigan is significantly higher than the proportion of deaths we would expect based on population representation alone (p<0.001).

Black individuals who died from COVID-19 were significantly younger (median [IQR]: 72 [63, 81], p<0.001) than White individuals (81 [72, 89]) and were less likely to be retired or unemployed (12.9%, p<0.001) than White decedents (22.2%) (Table 1). Among all COVID-19 deaths, 44.0% were either residents or employees of high-risk or congregate living facilities, such as long-term care and senior homes, homeless shelters, and prisons. Black decedents (29.7%) were less likely to have been living or working in high-risk or congregate living facilities than White individuals (54.8%, p<0.001).

**Table 1.** Demographic characteristics of individuals who died from COVID-19 in Michigan overall and by race. Data are median [IQR], n (%), or n/N (%), and p-values were calculated by Chi-Square tests or Kruskal-Wallis tests as appropriate. The study population overall includes deaths among individuals of all races, of which Black and White individuals make up 96.3%.

|  | Race |  |  | Overall<br>(n=6065) |
| --- | --- | --- | --- | --- |
|  | Black<br>n = 2341<br>(38.6%) | White<br>n = 3497<br>(57.7%) | p-value |  |
| **Sex** |  |  | 0.006 |  |
| Male | 1274 (54.4) | 1773 (50.7) |  | 3186 (52.5) |
| Female | 1067 (45.6) | 1724 (49.3) |  | 2879 (47.5) |
| **Age (median [IQR])** | 72 [63, 81] | 81 [72, 89] | <0.001 | 77 [68, 86] |
| **Congregate Living Facility Resident or Employee** | 695 (29.7) | 1915 (54.8) | <0.001 | 2666 (44.0) |
| **Retired or Unemployed** | 302 (12.9) | 776 (22.2) | <0.001 | 1128 (18.6) |
| **Number of Comorbidities** |  |  | <0.001 |  |
| None | 675 (28.8) | 566 (16.2) |  | 1293 (21.3) |
| One | 428 (18.3) | 636 (18.2) |  | 1116 (18.4) |
| Two or more | 1238 (52.9) | 2295 (65.6) |  | 3656 (60.3) |
| **Comorbidities** |  |  |  |  |
| Asthma or Reactive Airway Disease | 149 (6.4) | 170 (4.9) | 0.016 | 341 (5.6) |
| Cardiovascular Disease | 1235 (52.8) | 2293 (65.6) | <0.001 | 3647 (60.1) |
| Cancer | 191 (8.2) | 425 (12.2) | <0.001 | 639 (10.5) |
| Chronic Lung Disease | 369 (15.8) | 826 (23.6) | <0.001 | 1226 (20.2) |
| Diabetes Mellitus | 744 (31.8) | 978 (28.0) | 0.002 | 1819 (30.0) |
| Neurologic Disease | 460 (19.6) | 1242 (35.5) | <0.001 | 1741 (28.7) |
| Chronic Liver Disease | 64 (2.7) | 76 (2.2) | 0.199 | 149 (2.5) |
| Chronic Renal Disease | 498 (21.3) | 637 (18.2) | 0.004 | 1181 (19.5) |
| Other Immunosuppressive Conditions | 134 (5.7) | 204 (5.8) | 0.906 | 348 (5.7) |
| Other Chronic Diseases | 215 (9.2) | 505 (14.4) | <0.001 | 741 (12.2) |
| **Relevant Comorbidity Combinations** |  |  |  |  |
| Diabetes Mellitus, Cardiovascular, and Chronic Renal Disease | 214 (9.1) | 244 (7.0) | 0.003 | 488 (8.0) |
| Diabetes Mellitus and Cardiovascular Disease | 576 (24.6) | 828 (23.7) | 0.435 | 1480 (24.4) |
| Cardiovascular and Chronic Lung Diseases | 281 (12.0) | 660 (18.9) | <0.001 | 964 (15.9) |
| Cardiovascular and Neurologic Disease | 353 (15.1) | 935 (26.7) | <0.001 | 1312 (21.6) |

The most common comorbidity dyads among all COVID-19 deaths were diabetes and cardiovascular disease (24.4%), and cardiovascular and neurologic disease (21.6%). Black decedents had significantly higher rates of reporting asthma (p = 0.016), diabetes (p = 0.002), and chronic renal disease (p = 0.004) than White individuals who died from COVID-19 (Table 1), but significantly lower rates of cardiovascular disease (p<0.001), cancer (p<0.001), chronic lung disease (p<0.001), and neurologic disease (p<0.001). Additionally, Black decedents were more likely to have the combination of diabetes mellitus, cardiovascular disease, and chronic renal disease (p = 0.003) than White decedents, and less likely to have the combinations of cardiovascular and chronic lung disease (p<0.001) and cardiovascular and neurologic disease (p<0.001).

### Mortality Rates

Between March 16 to October 26, 2020, the COVID-19 mortality rate in Michigan was 5.4 per 10,000 population. Stratifying by race, the mortality rate was 3.6 times higher for Black populations (15.6 per 10,000 population) than White populations (4.3 per 10,000 population). Stratifying by age, the mortality rate for Black individuals under 40 years of age (0.50 per 10,000 population) is 7.4 times that for White individuals in the same age group (0.07 per 10,000 population). Among those aged 40 to 69 years, the mortality rate for Black individuals (18.4 per 10,000) was 8.5 times that of White individuals (2.2 per 10,000). Mortality risk for both races increased with age, where Black individuals aged 70 years and older had a mortality rate of 121.1 per 10,000 population and White individuals had a mortality rate of 27.7 per 10,000 population.

While the mortality rate in the White population did not vary substantially by sex (4.5 per 10,000 population for males and 4.2 per 10,000 for females), Black males have a significantly greater (31.2%) mortality rate than their female counterparts. Comparing males and females under the age of 40, the Black male mortality rate (0.77 per 10,000 population) is 10.8 times that of White males (0.07 per 10,000 population), and the Black female mortality rate (0.23 per 10,000 population) was 3.56 times that of White females in the same age group (0.06 per 10,000 population).

When stratified by age, sex, and number of comorbidities, the mortality rate for the Black population was significantly higher than that for the White population for every pair-wise comparison (p<0.001). Black males under the age of 65 years with no comorbidities had a COVID-19 mortality rate of 6.2 per 10,000 population, while White males with no comorbidities in the same age group experienced a lower mortality rate of 0.53 per 10,000 (Figure 1). Among those aged 65 and older, the COVID-19 mortality rate was 367.5 and 363.1 per 10,000 population for Black males and females respectively who had no comorbidities, compared to 31.6 and 35.9 per 10,000 among White males and females with no comorbidities. The relative difference in mortality rate is also stark when comparing Black and White individuals with comorbidities. Black males aged 65 and older with multiple comorbidities had a mortality rate that was 3.5 times that of White males with multiple comorbidities.

**Fig 1:**
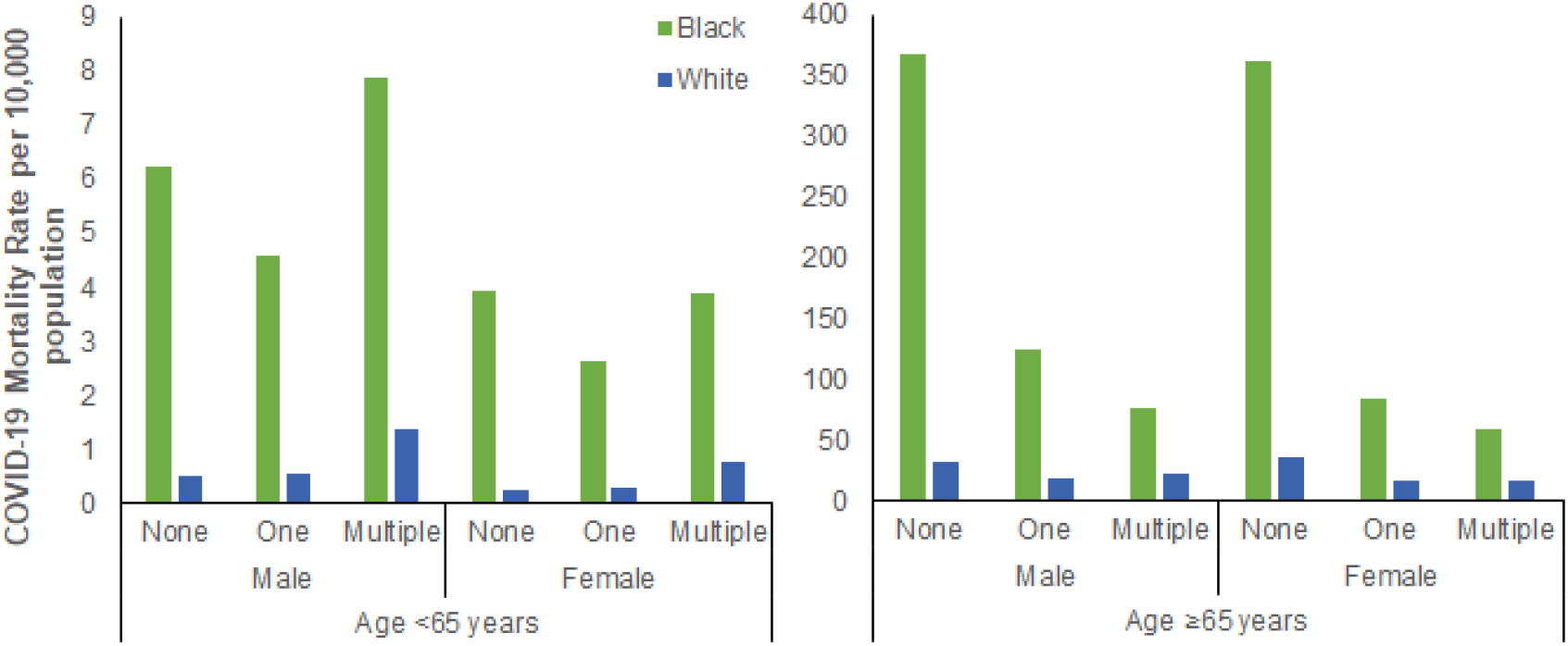
COVID-19 mortality per 10,000 population in Michigan by age, sex, number of comorbidities, and race. The population was first stratified by individuals under 65 years (left) and 65 years and older (right) then by sex, number of comorbidities, and race (White and Black). These mortality rates are based on 6,065 COVID-19 deaths that occurred in the state of Michigan between March 16 and October 26, 2020. Differences between the mortality rate of Black and White populations are statistically significant for every comparison.

### Spatial Analysis

Across the 198 ZIP codes in which at least one Black and at least one White individual died from COVID-19, the median mortality rate for Black populations was 167.9 per 100,000 [IQR: 89.4-251.8] compared to the median mortality rate of 64.0 per 100,000 population [IQR: 34.5-97.0] among white populations (Figure 2).

**Fig 2:**
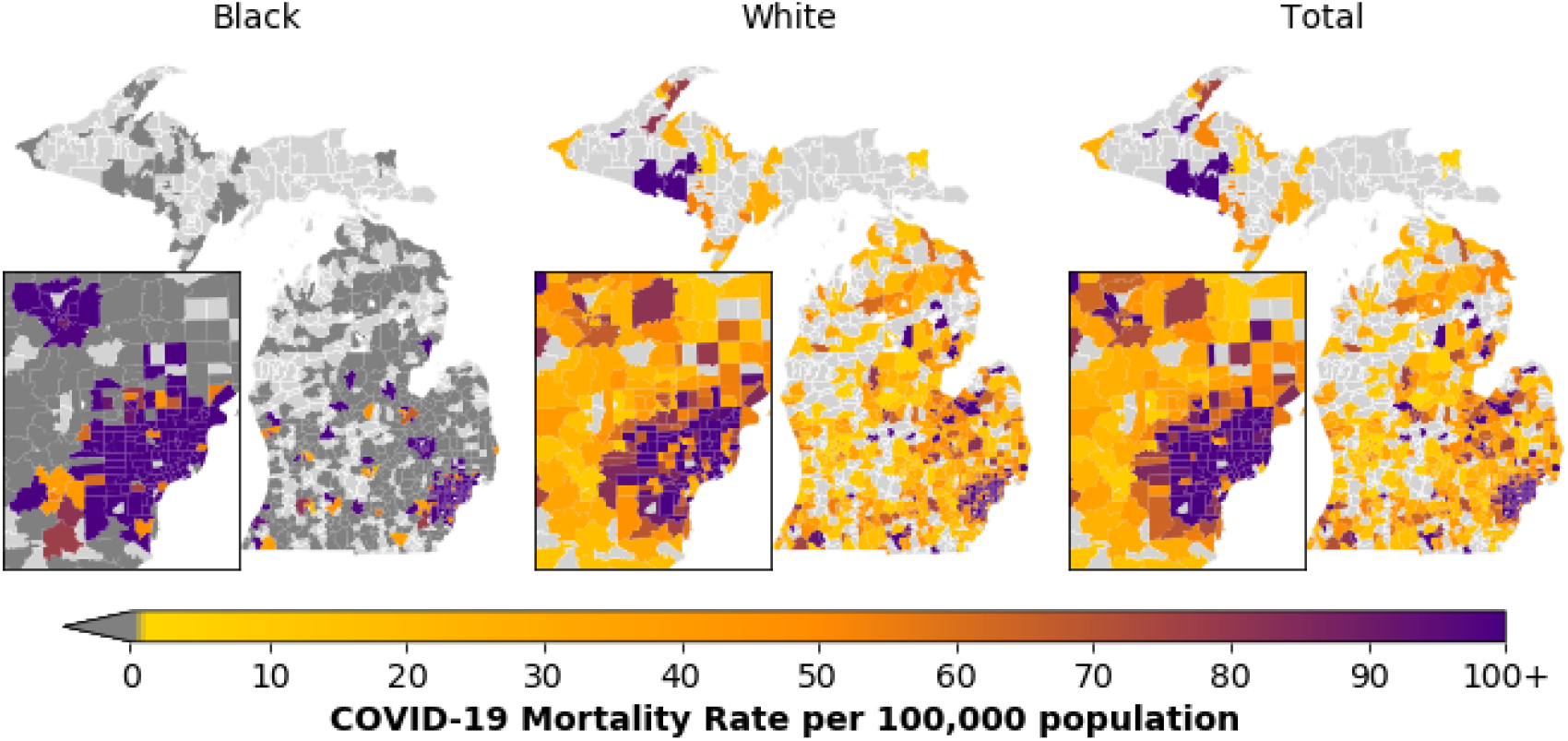
COVID-19 Mortality rates per 100,000 population among Black and White Michigan residents and Michigan residents overall by ZIP Code Tabulation Area. Mortality rate per 100,000 population ranges from 0 (yellow) to 100+ (purple). The highest mortality rate per 100,000 population is 5263. Dark grey regions indicate ZIP code tabulation areas where no COVID-19 deaths for a particular race occurred and light grey regions indicate ZIP code tabulation areas where no COVID-19 related deaths took place. These mortality rates are based on 6027 COVID-19 deaths among Michigan residents spread across the state between March 16 and October 26, 2020, of whom 5809 individuals are either White or Black. Total includes individuals of all races. The inset map represents the Detroit Metropolitan Area and Flint.

We found that mortality rates were higher among Black individuals in 84.8% of these 198 ZIP codes (Figure 2, Figure 3). For those ZIP codes in which individuals of both races died, mortality rates were significantly higher among Black individuals, as determined by Wilcoxon signed-rank test (p<0.001). In these ZIP codes, Black individuals had a median of 2.3 times [IQR:1.3-4.8] greater odds of dying from COVID-19 than White individuals (Figure 3). Detroit and Flint have the largest Black populations in Michigan and were both devastated by COVID-19 mortality.

**Fig 3:**
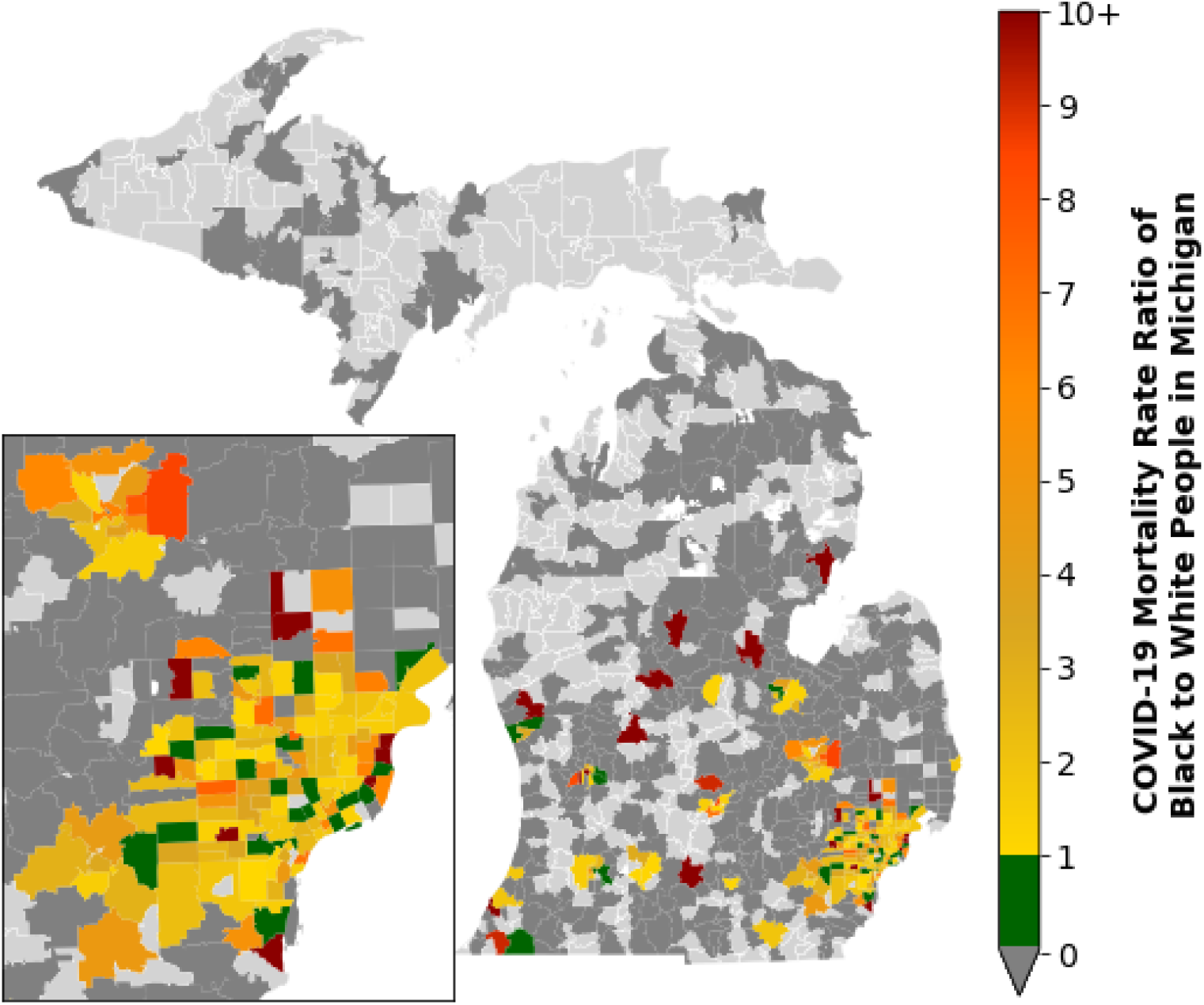
Mortality rate ratio for Black to White populations at the ZIP Code Tabulation Area level of residence in Michigan. There were 30 ZIP codes where deaths among Black individuals occurred, yet the White COVID-19 mortality rate exceeded the Black mortality rate (green). The Black COVID-19 mortality rate exceeds White mortality rate in 178 ZIP codes (yellow to red). Dark grey regions indicate ZIP codes where COVID-19 deaths occurred only among one race, Black or White. Light grey regions indicate ZIP code tabulation areas where no COVID-19 related deaths took place. The inset map represents the Detroit Metropolitan Area and Flint. These mortality rates are based on 5809 COVID-19 deaths among Black and White Michigan residents spread across the state between March 16 and October 26, 2020.

### Date of Symptom Onset, Hospitalization, and Death

The transmission intensity driven by exposure risk within a population is reflected by the timing of the peaks in symptom onset and mortality. Among all COVID-19 decedents in Michigan, dates of symptom onset and death were earlier for Black populations in comparison to White populations (Figure 4), indicating a steeper escalation in risk for Black individuals. The peak in symptom onset for Black individuals in Michigan was 6 days earlier (March 26, 2020) than White individuals (April 1, 2020). The median onset date and median date of death were 13 days earlier for Black individuals who died than White, and peak daily deaths occurred 3 days earlier for Black individuals than White (April 8, 2020 vs April 11, 2020).

**Fig 4:**
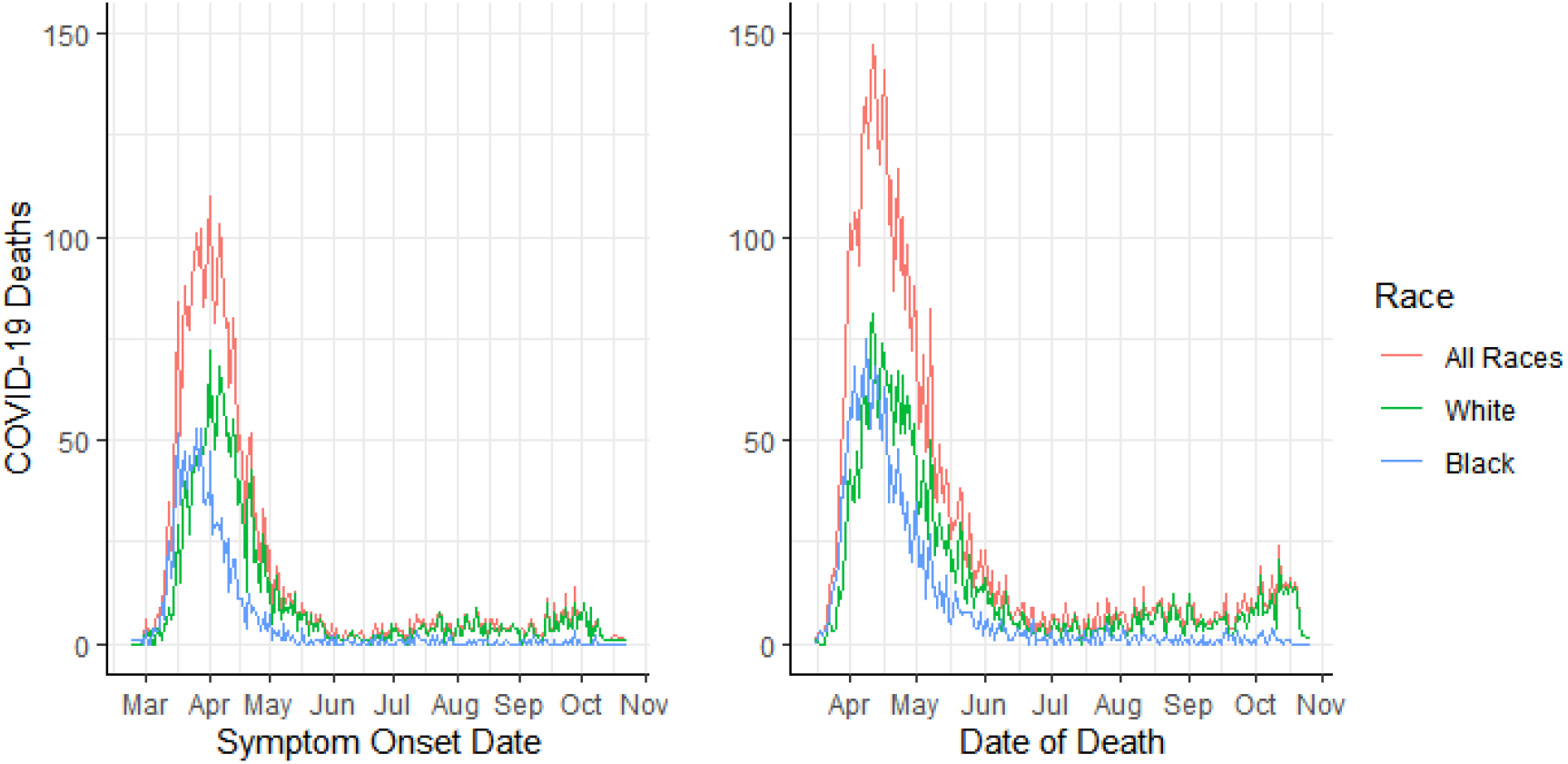
Time series of symptom onset and date of death among individuals who died from COVID-19 in Michigan by race. Date of symptom onset (n=4110) and death (n=6065) are displayed from February 23 to October 23, 2020 and March 16 to October 26, 2020, respectively.

Prompt medical attention is an important component of recovery. The time between symptom onset and hospitalization was a median of 1 day longer for Black decedents compared to White (p=0.020), after adjusting for number of comorbidities, age group, and sex.

## Discussion

Our highly granular analyses demonstrate that at the individual, ZIP Code and state levels, racial disparities in the burden and impact of COVID-19 are salient and reflective of pervasive inequities. By accounting for pre-existing comorbidities, we show that these underlying health conditions alone do not explain the racial disparities in COVID-19 mortality. In fact, when adjusting for age and health status, the gap becomes even more striking. Overall, Black populations in Michigan have 3.6 times the mortality rate of White populations. When subset to individuals under 65 years old and without comorbidities, Black individuals have 12.6 times the mortality rate of White individuals.

The racial disparity in mortality rate is most pronounced at younger ages. The significantly younger median age of Black deaths (72 vs. 81 years) results in even more years of life lost in this community than the differential mortality rate alone would suggest. High mortality rates observed in young Black populations may be highlighting the vulnerabilities of individuals unable to participate in stay-at-home orders during the pandemic. For instance, the flexibility to work from home is often linked to higher income and to industries in which Black individuals are underrepresented. While 47 to 49% of White individuals report being able to work from home, only 34 to 39% of Black individuals have the same privilege.^28^ Furthermore, Black Americans are disproportionately employed in low-wage and high-contact essential service industries ^29^ within which sick leave is often discouraged and uncompensated. Compounding these issues, low-wage earners have more children ^30^ with an average of 2.4 dependent family members.^31^ In the absence of sick leave benefits under these circumstances, infectious individuals in essential service industries are more likely to spread COVID-19 to their disproportionately Black coworkers.^29^ Our finding that peak incidence occurred six days earlier among Black individuals further evidences heightened exposure of Black individuals.

Our results regarding the duration between symptom onset and hospitalization indicate that Black COVID-19 patients do not receive medical attention as promptly as their White counterparts, a factor which is known to influence survival.^32^ Three non-exclusive factors may contribute to this difference: financial barriers deterring care-seeking,^33^ test scarcity, and racial bias among healthcare providers.^34^ In the US, Black populations represent 12.3% of the total employed population, and make up 25% of those employed in industries experiencing the highest proportion of job losses due to COVID-19.^21,35^ This disproportionate burden of unemployment the Black community, and the precarious linkage of health insurance to employment in this country, may be serving as a deterrent to care-seeking when COVID-19 symptoms arise. Upon seeking medical attention, those living in densely populated inner cities have faced inadequate access to testing.^36^ These issues interplay with the racial composition of an area. Detroit, the most populous city in Michigan, is also home to a larger proportion of Black individuals than any other city in the United States. Additionally, the SARS-CoV-2 test used in Detroit during the early months of the pandemic was found to miss 45% of positive cases.^37,38^ Patients receiving these false negative results are likely to have received delayed or inappropriate care, compounding the other factors described above. Finally, physician racial biases may affect patient health outcomes.^34,39^

Racial disparities in access to healthcare, educational opportunities and economic security predate the COVID-19 epidemic. At least 90% of the ZIP codes within the predominantly Black cities of Detroit and Flint have rates of child poverty that are higher than the national average.^40^ The anti-Black systemic racism in our political system is paralleled by the racialized nature of COVID-19.^29^ Inadequate governance nationwide has hindered pandemic response, analogous to the ongoing mishandling of the Flint water crisis that has arisen from a series of misguided governmental, social, and economic policies.^41^ The effects of COVID-19 shown here highlight a need for corrective resource allocation by federal and local governments that would mitigate the toll of public health crises on vulnerable populations. Universal healthcare, living wages for all workers, and paid sick leave will be important first steps in addressing racial inequities in the US.

As the COVID-19 pandemic continues to unfold, it is imperative to ameliorate disparities as both a pandemic response and prevention measure. Systemic disenfranchisement of Black individuals in the US underlies circumstances leading to racial disparities in COVID-19 exposure, the timeliness of treatment and case fatality rates.^29^ While structural racism as an infectious disease risk factor is increasingly being recognized,^42^ the US response to this pandemic demonstrates our society’s continued negligence toward the wellbeing of all Americans. It is not only the longstanding increased risk of comorbidities in the Black population that drives racial disparities in COVID-19 mortality, but socio-political and economic factors that impact COVID-19 exposure and medical care. Rectifying these inequities is urgent both to limit the devastation of COVID-19 and to protect against future public health crises.

## Data Availability

Aggregated data are provided in the manuscript. Raw data may be requested from the Michigan Department of Health and Human Services.

## Acknowledgements

We would like to thank Seth Eckel and Adam Hart for linking Vital Records and MDSS data, and to remember all those in Michigan who lost their lives to COVID-19.

