## Supplement for "Racial disparities in COVID-19 mortality across Michigan, United States"

Section 1

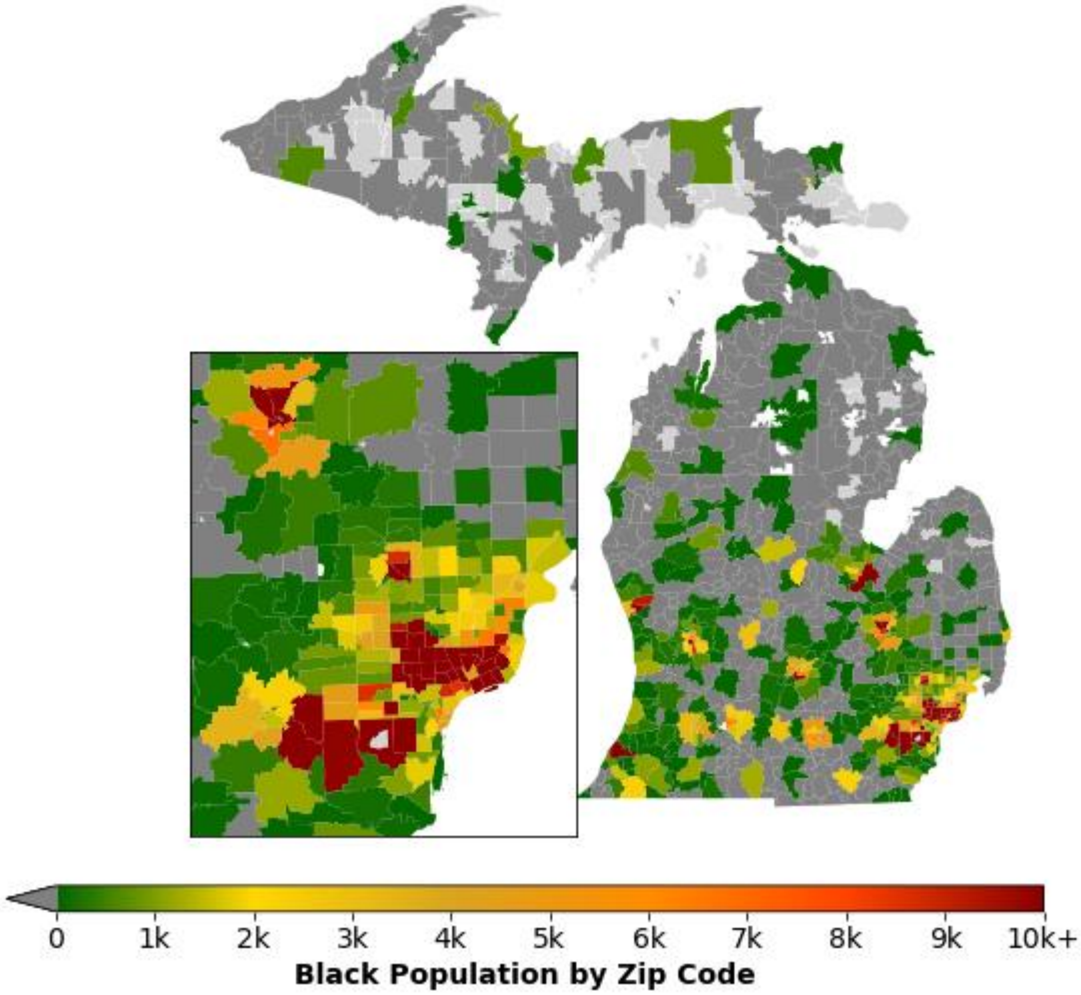


**Fig S1: Black population by Zip Code Tabulation Area in Michigan.** Dark Grey represents ZIP codes with no black individuals and light grey indicates ZIP codes where no race-based population data was available.

**Table S1: COVID-19 Deaths in Michigan by age group, sex and race from March 16 to October 26, 2020.**

| | Black (n=2341) | | White (n=3497) | | All Races (n=6065) | |
|---|---|---|---|---|---|---|
| **Age Group** | **Male** | **Female** | **Male** | **Female** | **Male** | **Female** |
| <40 | 34 | 10 | 14 | 12 | 50 | 24 |
| 40-49 | 65 | 45 | 32 | 21 | 110 | 70 |
| 50-59 | 172 | 93 | 112 | 60 | 295 | 160 |
| 60-69 | 313 | 226 | 296 | 169 | 646 | 404 |
| 70-79 | 389 | 310 | 498 | 381 | 931 | 720 |
| 80+ | 301 | 383 | 821 | 1081 | 1154 | 1501 |

**Table S2: Residents/Workers at High Risk or Congregate Living Facilities**

| | | | Deaths among Residents of and Workers at High Risk or Congregate Living Facilities | |
|---|---|---|---|---|
| | | | Race | |
| **Age (years)** | **Sex** | **Number of Comorbidities** | **Black** | **White** |
| <65 | Male | 0 | 13 | 8 |
| | | 1 | 16 | 12 |
| | | 2+ | 39 | 54 |
| | Female | 0 | 7 | 2 |
| | | 1 | 5 | 7 |
| | | 2+ | 34 | 47 |
| 65+ | Male | 0 | 54 | 77 |
| | | 1 | 45 | 102 |
| | | 2+ | 165 | 560 |
| | Female | 0 | 54 | 113 |
| | | 1 | 46 | 174 |
| | | 2+ | 217 | 759 |

* High Risk or Congregate Living Facilities include: long-term care homes, skilled nursing facilities, assisted living facilities, homeless shelters, federal prisons, Michigan Department of Corrections prisons, county jail, juvenile justice facilities, foster care, and others, including senior, retirement, and group homes.

**Table S3: Deaths by Race in Michigan.** Deaths are presented by race that occurred in Michigan from March 16 and October 26, 2020. For racial groups that reported fewer than 5 deaths, we have aggregated deaths.

| **Race** | **Deaths (n=6065)** |
|---|---|
| White | 3497 |
| Black | 2341 |
| American Indian | 18 |
| Asian Indian | 28 |
| Chinese | 7 |
| Filipino | 22 |
| Japanese, Korean, or Vietnamese | 7 |
| Other Asian Race | 30 |
| Other or Unknown | 115 |

Section 2

**Table S4. Calculating Proportion of population that has no comorbidities and 1 comorbidity.** Using data from the National Health Interview Survey on the percentage of adults with one or more and two or more chronic conditions by age group and sex,[43] we were able to calculate the percentage of individuals in each age group and sex with no comorbidities and with only one comorbidity. For individuals aged 65 and older, we used this information in combination with Medicare Beneficiary data on those 65 years and older from Michigan on the proportion of beneficiaries with 0 or 1 comorbidity and 2 or more comorbidities by race. We assumed the distribution of comorbidities for each sex do not vary by race and we created estimates for Michigan comorbidity distribution by age, sex, and race. For individuals under age 65 years, we calculated race-specific proportions by applying Medicare data by race to the National Health Interview Survey data by sex and number of comorbidities. Medicare data for the under 65 population was not used as the primary source for comorbidity burden distribution due to strict enrollment criteria in this age group (disability, end-stage renal disease, and Amyotropic Lateral Sclerosis).[44]

| | | **Number of Comorbidities** | | | |
|---|---|---|---|---|---|
| **Sex** | **Age Group (years)** | 0 | 1 | ≥1[43] | ≥2[43] |
| Male | 55 to 64 | 32.3% | 35.4% | 67.7% | 32.3 |
| | ≥65 | 17.0% | 31.6% | 83.0% | 51.4 |
| Female | 55 to 64 | 28.9% | 29.6% | 71.1% | 41.5 |
| | ≥65 | 12.4% | 28.2% | 87.6% | 59.4 |

Section 3:

**Related causes of death and comorbidities included under the categories of:**

*Other Chronic Conditions:*
Anemia, Chronic venous thromboembolism (deep vein thrombosis and pulmonary embolism), Bipolar disorder, Osteoarthritis, Depression, Osteoporosis & osteopenia, Alcohol Abuse, Fragile X syndrome, Hemochromatosis, Hepatitis C & Hepatitis B, Schizophrenia, Addison's Disease, Chronic pancreatitis, Chronic Wounds, Sickle Cell Disease.

*Other Immunosuppressive Conditions:*
Rheumatoid arthritis, Bullous Pemphigoid, HIV, Polymyalgia rheumatica, Inflammatory bowel disease (Crohn's disease & Ulcerative Colitis), Sarcoidosis, IgG4 Disease, Systemic Lupus Erythematosus, Giant Cell Arteritis (or Temporal arteritis), IgA Deficiency, Psoriatic arthritis, Solid Organ Transplant.
